# Impact of the household environment risk for maintenance of natural foci of *Leishmania infantum* transmission to human and animal hosts in endemic areas for visceral leishmaniasis in Sao Paulo State, Brazil

**DOI:** 10.1101/2021.05.18.21257380

**Authors:** Patricia Sayuri Silvestre Matsumoto, Roberto Mitsuyoshi Hiramoto, Virgínia Bodelão Richini Pereira, Valéria Medina Camprigher, Helena Hilomi Taniguchi, José Eduardo de Raeffray Barbosa, Luiz Ricardo Paes de Barros Cortez, Elivelton da Silva Fonseca, Raul Borges Guimarães, José Eduardo Tolezano

**Affiliations:** Parasitology and Mycology Center, Adolfo Lutz Institute (IAL), Sao Paulo, Sao Paulo, Brazil; Adolfo Lutz Institute, Regional Laboratories Center II Bauru, Bauru, Sao Paulo, Brazil; Center for Zoonoses Control of Bauru. Health Secretariat of Bauru; Bioterium nucleos, Adolfo Lutz Institute (IAL), Sao Paulo, Sao Paulo, Brazil; Federal University of Uberlândia, Brazil; Department of Geography, Sao Paulo State University/Faculty of Sciences and Technology (FCT/UNESP), Presidente Prudente, Sao Paulo, Brazil

**Keywords:** Vector-borne diseases, disease surveillance and response, visceral leishmaniasis, dogs, spatial modeling, kernel density, geostatistics, generalized additive model, high-risk, public health

## Abstract

When it comes to visceral leishmaniasis (VL) in Brazil, one of the main targets of public health policies of surveillance is the control of domestic canine reservoirs of *Leishmania infantum*. This paper aims to evaluate the effect of the household environment risk in the maintenance of natural foci and in the transmission to human and animal hosts in an endemic city for VL, Bauru, in Brazil. We collected 6,578 blood samples of dogs living in 3,916 households from Nov.2019 to Mar.2020 and applied geospatial models to predict the disease risk based on the canine population. We used Kernel density estimation, cluster analysis, geostatistics and Generalized Additive Models (GAM). To validate our models, we used cross-validation and created a ROC graph. We found an overall canine VL (CVL) prevalence of 5.6%. Odds ratios (OR) for CVL increased progressively according to the number of canines for >2 dogs (OR 2.70); households that already had CVL in the past increased the chances for CVL currently (OR 2.73); and the cases of CVL increase the chances for human VL cases (OR 1.16). Our models were statistically significant and demonstrated an association between the canine and human disease, mainly in VL foci that remain endemic. Although the Kernel ratio map had the best performance (AUC=82), all the models showed high risk in the city’s northwest area. Canine population dynamics must be considered in public policies and geospatial methods may help target priority areas and planning VL surveillance in low and middle-income countries.

**Highlights:**

- Two or more dogs in a household increase the chances for canine visceral leishmaniasis.
- Canine visceral leishmaniasis or households with positive dogs increase the chances for human visceral leishmaniasis.
- Households that already had an infected dog increase the chances for canine visceral leishmaniasis, and it can work as silent endemic areas.
- More than 40 dogs in an area of influence of household (100m buffer) increase the chances for canine and human visceral leishmaniasis.
- Canine population dynamics must be considered in public policies regarding visceral leishmaniasis control in low and middle-income countries.
- Spatial analysis tools can bring new insights into decision-making and public policies regarding visceral leishmaniasis.

## 1. Introduction

Leishmaniasis is a group of infectious diseases caused by a protozoan of the *Leishmania* genus that affects humans and animals. The transmission occurs by the bite of the dipterous of the subfamily Phlebotominae, the sand flies. It is considered one of the most widely distributed neglected diseases worldwide(1), being a health problem in North and East Africa, West and East Asia, and the Americas(2). More than one billion and a half persons live in risk areas for leishmaniasis around the world. In 2015, between 50 and 90 thousand new cases were estimated per year, with an incidence rate of 2.27 per 100 thousand inhabitants. Only six countries, including Brazil, accounted for about 90% of all new cases each year(3)(4). From 2005 to 2019, Brazil registered 51,931 cases. In the last update in 2021, Brazil notified a mean of 3,462 human cases of visceral leishmaniasis (VL) in the last 15 years(2).

VL is a focal disease and its epidemiology differs according to nosogeography entity, which means that different spatial patterns for each species of *Leishmania* occur, and different strategies for the control of leishmaniasis are demanded. In Latin America, the main reservoir is the domestic dog. However, for control programs, an integrated knowledge about the ecological niche of the vector and environmental conditions is fundamental to address effective measures(5). For this reason, geospatial modeling is a valuable instrument to target interventions of control programs(6).

In Brazil, there is a great difficulty for the effective implementation and operation of the VL control programs (7). Overall, the Brazilian Visceral Leishmaniasis Control Program (VLCP) is based on the control of canine reservoirs, which consists in serosurvey and culling of dogs; control of the vector spraying insecticides inside the households; and early diagnosis and treatment of human cases (8).

The first evidence of VL in Sao Paulo state was the presence of *Lutzomyia longipalpis* in the urban area of Araçatuba municipality in 1997 (9). In 1998, autochthonous VL dogs were reported, and for the first time in the state, the sand fly was suspected as the vector; and in 1999 autochthonous human cases were reported for the first time (10)(11).

Several factors may be responsible for increasing the cases and the number of deaths in Sao Paulo state, such as difficulty for early diagnosis and specific treatment in human; difficulty in the correct identification and control of domestic reservoirs; and difficulty in controlling the vector population (8). In addition, there is unclear knowledge about other determinants that may influence the design of novel strategies for control and prevention of VL (12).

In Sao Paulo state, Bauru had the first evidence of VL in 2002 when the sandflies were found and the first autochthonous infection in a dog was reported. The first human records occurred in 2003. Since that time, there have been 580 cases and 46 deaths, a lethality rate of 7.9% from 2003 to 2020(13). The municipality of Bauru was chosen to perform this research because of the high number of cases and its endemicity in the region. In Bauru, there is a lack of information about the spatial distribution and a long-term follow-up of CVL, information that could aid in the global understanding of the problem. The spread of the disease (human cases) came from one cluster in the west to east, and the environmental characteristics suggest a causal relationship of deforestation and human occupation is associated with the emergence of new VL cases (14). Mapping the exact occurrence of the human or canine cases may help better understand the disease and plan public policies regarding VL.

This study aimed to calculate the impact of the household environment risk for visceral leishmaniasis using geospatial methods. We hypothesize that: a) the number of dogs in the households tends to increase human and/or canine VL cases. b) the urban area is stratified by different geographical profiles that allow the remaining endemicity, needing targeted strategies as control measures. Using spatial analysis and statistical approaches, we constructed a space framework based on a large serosurvey conducted between November 2019 and March 2020 in the urban area of Bauru. Of note, there is no existent study using a machine learning-based approach of CVL risk prediction considering the number of dogs. Spatial analysis may be useful to specific interventions to understand VL better.

## 2. Materials and Methods

Bauru is a central municipality of Sao Paulo state (22°18’52” S, 49°03’31” W), crossed by important highways: SP-300 - Marechal Cândido Rondon Highway, SP-294 - Comandante João Ribeiro de Barros Highway, SP-321 - Cezário José de Castilho Highway, and SP-225 - Engenheiro João Batista Cabral Rennó Highway, giving access to the countryside cities of the state as well to the capital, Sao Paulo.

According to the Köppen-Geiger climate classification updated system (15)(16), Bauru climate is classified as Cfa, which means temperate, without dry season, and with hot Summer. The soil is unsaturated, reddish and dark brown, fine clay sand texture, underlain by sandstone of the Bauru group. The urban areas vegetation is Tropical Semideciduous Forest mixed with Cerrado, highly impacted by urbanization variable patchy pattern. The average altitude of Bauru is 527.4 m.

The population comprises an estimative of 379,297 inhabitants (IBGE, 2020). A research was conducted in 20,958 households in the Sao Paulo state countryside, in which 52.6 % proven dog ownership, with an average of 1.6 dogs at home and an inhabitant ratio of 1:4 dogs per person(18). Following this study, we estimated the dog population at 99,815, according to the Brazilian Geographic and Statistics Institute (17).

### 2.1. Data sources and Laboratory Diagnostic Tests

The study analyzed 6,578 samples and surveyed the dog owners from November 2019 to March 2020. Dual-Path Platform rapid test (TR-DPP, Biomanguinhos®, Rio de Janeiro, Brazil,) is used by the Brazilian VLCP to test the samples. The TR-DPP® is a test for *Leishmania infantum* based on the reaction of IgG to the antigen K28. Enzyme Linked Immunosorbent Assay ELISA - Biomanguinhos® is used to confirm the diagnoses in the VLCP. It is the reaction of soluble and purified antigens of *Leishmania* promastigotes, obtained from cultures and adsorbed in microtiter wells with *Leishmania*-specific antibodies present in serum samples. The diagnostic was run in a Multiscan spectrophotometer using a 450 nm filter and cutoff values (“Cutoff” = CO): CO = average negative controls x 2. The diagnoses were performed according to the manufacturer’s instructions and the directions of the VLCP.

### 2.2. Definition of cases

A combination of TR DPP® and ELISA reagent was considered a positive result according to the Brazilian VLCP recommendations for canine diagnoses, routinely used by the Centers for zoonoses control in Sao Paulo (19). TR DPP® non-reagent was considered a negative result - supplementary material. The prevalence was calculated based on the outcome, being a proportion of a dog found positive for CVL divided by the analyzed dog population. The consent for sample collection involving domestic dogs was provided by the dog owners in the areas surveyed. All serosurvey was supervised by the veterinary group of the Adolfo Lutz Institute in conjunction with the veterinarians responsible for the Center for Zoonoses Control in Bauru municipality. Households without dogs, closed or that refused to give the consent were excluded from the analysis.

The human laboratory diagnoses are based mainly on serological methods and microscopic diagnoses (parasitological). When amastigotes are identified, it is considered a certainty diagnostic. Patients with clinical manifestation and reagent rapid immunochromatographic test rK39 and/or Indirect immunofluorescence with titers equal to or greater than 80 are considered positive for VL (19). Human cases addresses come from the epidemiological surveillance center (CVE) (13).

### 2.3. Study design

Canine data were grouped into the households. The number of dogs was categorized as binary data to verify each risk group: one dog; two dogs; and more than two dogs. Households that already had a positive dog or a human case were also categorized as binary. We considered 1 for cases and 0 for non-cases of VL.

#### 2.3.1. Mapping

The addresses of dogs and households were geocoded by an application programming interface (API) of Google Maps (Google®), based on the municipality’s cartographic street map. A lower score of geocoding was topologically adjusted to ensure the correction of georeferencing. Point features were plotted in a Geographic Information Systems (GIS) ArcGIS 10.2.2 (ESRI, Imagem). Points (households) were categorized as negative or positive for VL in each survey. Figure 2 shows the mapped data.

**Figure 1:**
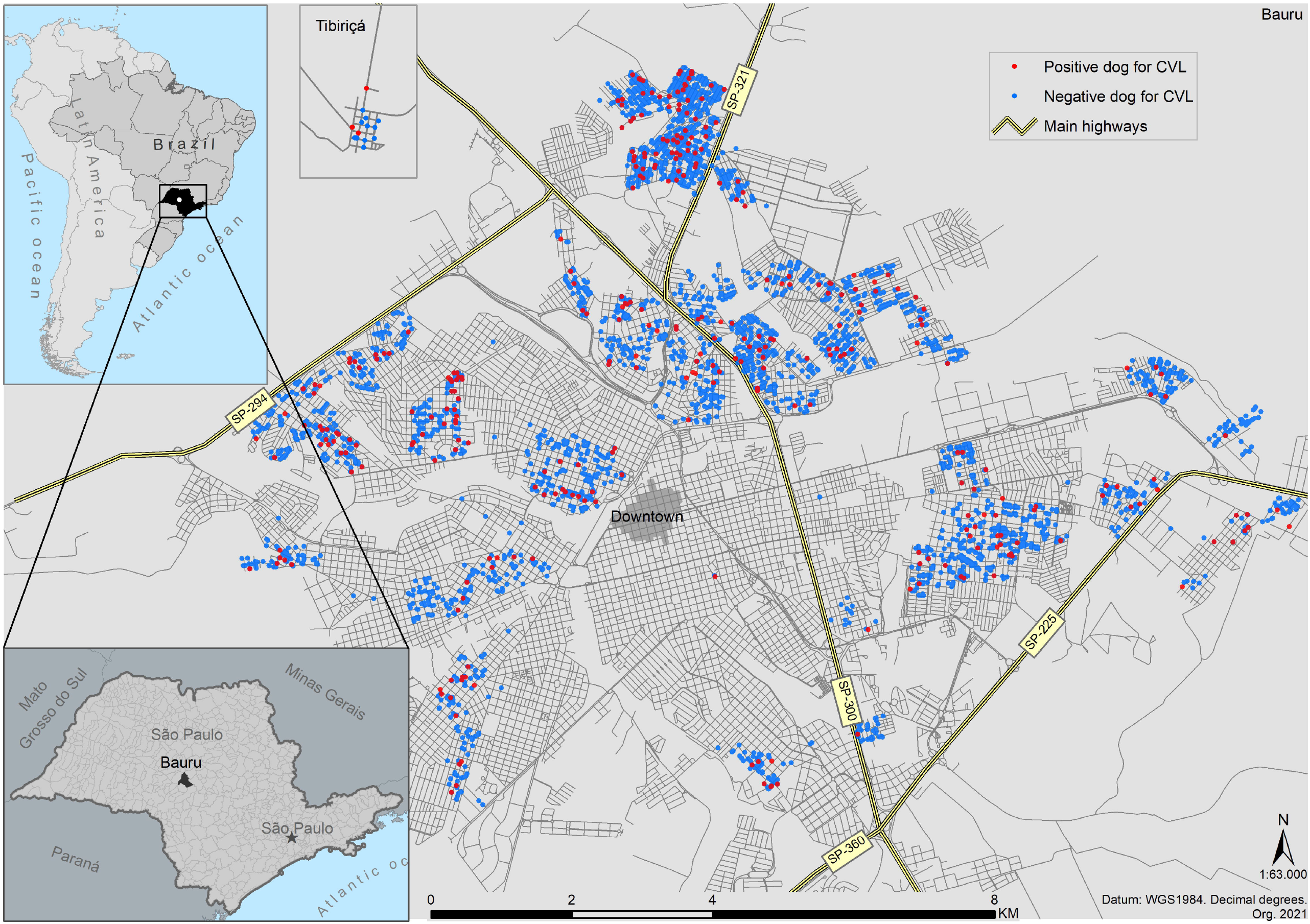
Geospatial location of South America, Brazil, Sao Paulo state, and the serosurvey in Bauru’s urban area. A total of 6,578 blood samples of dogs were analyzed. Points represent each dog’s address. Positive dogs for CVL are represented by red dots and negative by blue. Points are overlapped because of the spatial resolution of the cartographic scale.

**Figure 2.**
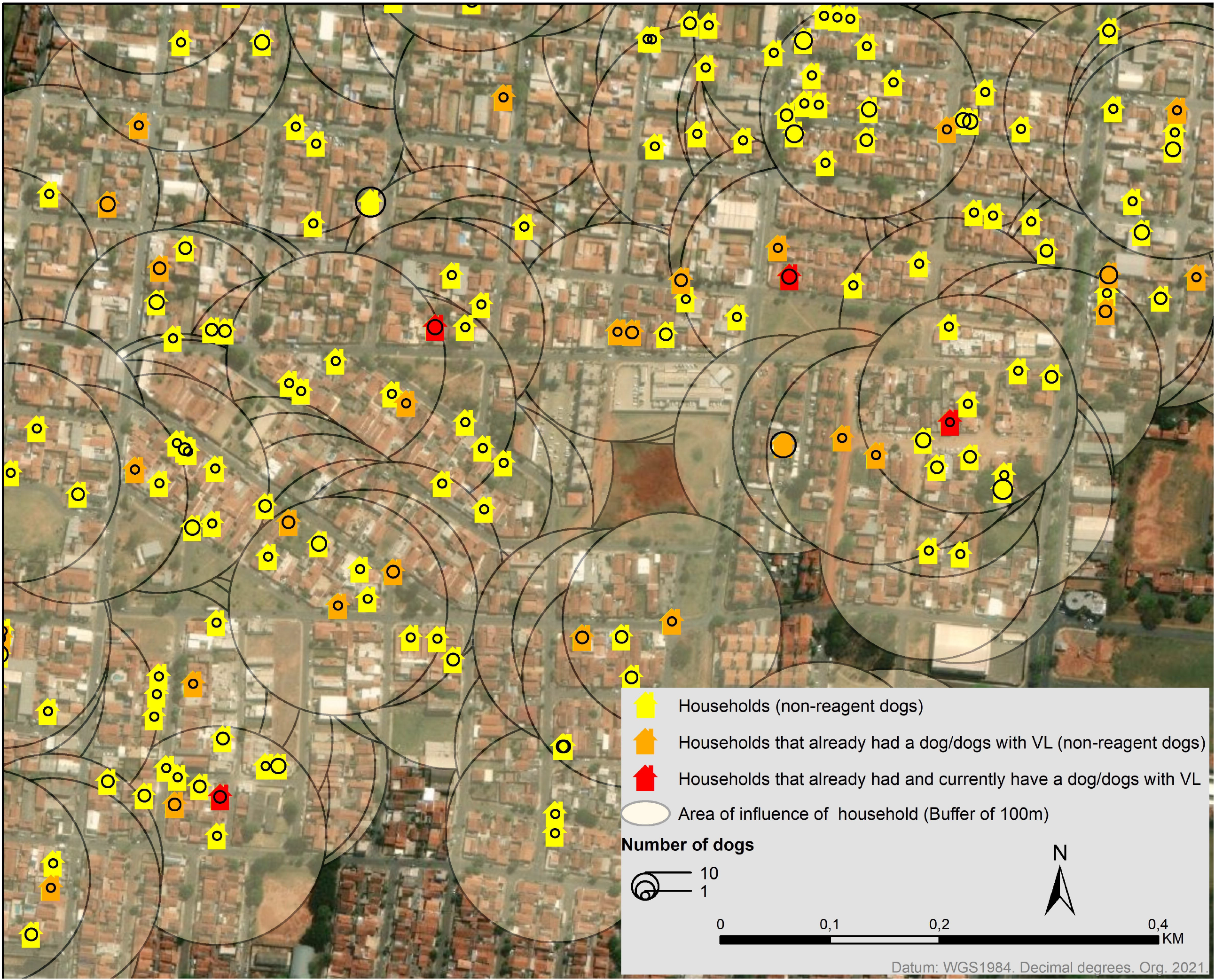
Geocoded households of Bauru, stratified by the presence of positive dogs and buffer zones. The households are identified according to the conducted surveys. Yellow, orange, or red symbols represent the dog’s households sampled. Yellow had not an infected dog; orange had an infected dog (in the past); red had and currently has an infected dog. Proportional circles represent the number of dogs in each household. We created a buffer of 100m in each sampled household to calculate the number of dogs, positive or negative dogs in this area.

To run the Generalized Additive Model (GAM), we created a fishnet grid of 1000×1000 of 50m containing the number of dogs per domicile in each coordinated. We calculated the number of dogs based on the human population by census tracts (20), in a ratio of 1:4 dogs/inhabitants (18). The mean number of dogs was divided by the mean number of households in each census tract. We then created a centroid and calculated the Inverse distance weighted (IDW) interpolation, considering the mean number of dogs per domicile. The grid values were extracted from the raster surface generated by the IDW - supplementary material.

To analyze the area of influence of households with infected dogs in the environment, we created buffer zones of 100m (Figure 2) – supplementary material. We then calculated the number of dogs, negative dogs, and positive dogs using spatial analysis tools. Finally, we aggregated features of point data into polygons, using the census tracts database, to stratify the prevalence spatially.

### 2.4. Statistical analysis

For all the performed calculations, we considered a significant value at p≤0.05. We used the geographic information system (GIS) ArcGIS 10.2.2 and R language, with several packages described in the sections below.

#### 2.4.1. Pearson’s correlation

Pearson’s correlation was calculated to identify a possible association between the number of cases of CVL and: i) the number of investigated samples or ii) the number of households that already had an infected dog, or iii) the number of households that already had and currently have an infected dog/dogs.

#### 2.4.2. Binary logistic regression

We tested if the households with an infected dog (outcome = 0 for a household with no infected dog/dogs or outcome = 1 for a household with an infected dog/dogs) or an area of influence of household (outcome = 0 for areas of influence of household with no infected dog/dogs or outcome = 1 for area of influence of household with infected dog/dogs) could possibly increase the chances to have cases of the disease.

#### 2.4.3. K-function

Being aware of spatial dependence of CVL promoting different risks or protection, we evaluate, locally, the spatial interactions in the urban neighborhoods. Ripley’s K-function with 999 permutations was applied to identify households’ spatial patterns at distances (21). In this function, K(t) is the number of events within a distance of an arbitrary event, divided by the overall density of events. We plotted maximum and minimal envelopes of the simulated values of K(t), giving the statistical significance for clustered or dispersed patterns – supplementary material.

#### 2.4.4. Cluster analysis

We used cluster analysis to detect significant concentrations of CVL within Confidence Intervals (CI) of 90%, 95%, and 99% - supplementary material. Clusters were calculated using Getis-Ord Gi statistic, which identifies features with either high or low values cluster spatially. The pattern can be expressed by clustered, dispersed, or random features and represents a measurable spatial aggregation unit (ESRI, Imagem).

#### 2.4.5. Kernel density

Using the K-funcion dependence, we choose the minimal distance of concentration of our data, 0.5 km, to set the bandwidth. We used the quartic kernel function (22), which is given by

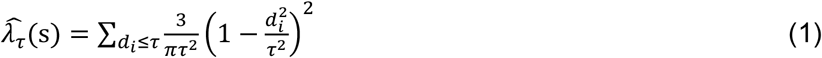

where:

i = *1,…,n* are the input points.

d_i_ is the distance between the point s and the observed event in location, s_i_ and τ is the radius centered on s.

We plotted Kernel density maps for CVL cases and canine samples. A Kernel density ratio map was then performed (CLV: samples), which gives a visualization of the risk for the disease.

#### 2.4.6. Geostatistical approach

According to the number of dogs, a geostatistical approach was performed to predict the higher risk areas for CVL. We used the Ordinary Kriging method and select two datasets: cases of CVL and number of dogs. We adjusted data in a stable model in a semivariogram, in which for a set of experimental values z(x) and Z (x1+*h*), separated by *h* distance, is defined by the equation 2:

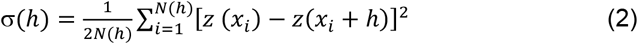

Where,

*N(h)* is the number of experimental pairs;

*h* is the regular interval that separates *z(xi)* e *z(x*_*i*_ *+h)*

Geostatistics parameters were adjusted as follow: number maximum and minimal neighbours= 5 and 2, respectively; lags=12; lag size=0.64; nugget=0.46; range=3.8974; sill=0.062 and 45 degrees – supplementary material.

#### 2.4.7. Generalized additive model

We run a GAM according to an approach reported for case-control data (23)(24), in which we considered Y_*i*_ = 1 (cases) and Y_*i*_ = 0 (non-cases), *d* is the number of dogs in location *i*, and P(Y_*i*_ = 1| d_*i*_ S_*i*_) is calculated according to equation 2:

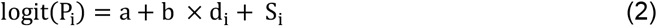

where,

a is the ratio of cases to non-cases,

b is the coefficient for the number of dogs per household,

S_i_ is a function of the residual spatial variation after accounting for the effect of the number of dogs.

We model S_i_ by a locally estimated scatterplot smoothing (LOESS) regression smoother against the Universal Transverse Mercator coordinates. We choose the optimal smoother parameter of the models based on Akaike’s Information Criterion (AIC)(25) after testing multiple bandwidths. We predicted the adjusted log odds for each location and omitted the covariate and smoothing terms through a null model. GAM was run in RStudio (4.0.0) using the ‘gam’ package, and the grid in ArcGIS 10.2.2.

#### 2.4.8. Cross-validation

For further analysis, we validated our data using cross-validation. We created random samples in ArcGIS and then split our database into training (75%, 2,937 points) and testing (25%, 979 points). Spatial models were created using the training dataset to predict the risk for the testing dataset. For each model, the best threshold was chosen, and we calculated specificity, sensitivity, and accuracy for correctly predicting the observed value of a case or non-case at the testing coordinates. To sum up, we calculated the area under the receiver operating characteristic (ROC) curve (AUC) with 95% confidence interval, which plots the true positive rate versus false positive rate, allowing identifying the performance of the models. We used the ‘pROC’ and ‘ggplot2’ packages in RStudio.

## 3. Results

The current study investigated 6,578 dogs (Table 1). Anti-*Leishmania* spp. antibodies were present in 8.1% of TR DPP® (535/6,578) and 5.6% in both TR DPP® and ELISA diagnoses (369/6,578). We found different spatial prevalence in the investigated census tract, ranging from 0 to 50%, but the mean prevalence was 2.67%. Higher prevalence (>7.5%) was regularly distributed in the city in the sampled area (Figure 2).

**Table 1:** CVL diagnostic, dog count in the households, and buffer zone extraction versus the number of dogs in Bauru.

| Category | Description | n | % |
| --- | --- | --- | --- |
| *Diagnoses* | Investigated dogs (samples) | 6,578 | 100 |
|  | Reagents samples (TR DPP®) | 535 | 8.1 |
|  | Non-reagents samples (TR DPP®) | 6,040 | 91.8 |
|  | Positive dogs (reagent for TR DPP® and ELISA) | 369 | 5.6 |
| *Households* | Investigated households | 3,916 | 100 |
|  | Households with only one dog | 2,269 | 57.9 |
|  | Households with two dogs | 1,084 | 27.6 |
|  | Households with more than two dogs | 563 | 14.3 |
|  | Households that already had a positive dog/dogs | 656 | 16.7 |
|  | Households with a positive dog/dogs currently | 341 | 8.7 |
|  | Households that already had a positive dog/dogs and currently have positive dog/dogs | 112 | 17.0 |
| *area of influence of household (buffer of 100m)* | > 0 ≤ 10 dogs | 1,402 | 35.8 |
|  | > 10 ≤ 20 dogs | 1,483 | 37.8 |
|  | > 20 ≤ 30 dogs | 576 | 14.7 |
|  | > 30 ≤ 40 dogs | 248 | 6.3 |
|  | > 40 ≤ 58 dogs | 185 | 4.7 |

Diagnoses summarize the serological diagnoses and the results for investigated dogs; households represent dog's address; the area of influence of household counts the dogs inside a buffer of 100m of radius.

### 3.1. Cluster analysis

We identified a clustered pattern of households with CVL with statistical significance from approximately 0.5 to 6.5 km and a clustered pattern of human cases from 0.5 to 4 km (*Figure 3S*). Spatially, we found clusters of high values (hot spots) (Figure 3) in west, north, east, south, northeast, southwest, southeast, and in Tibiriçá, a district of the municipality.

**Figure 3:**
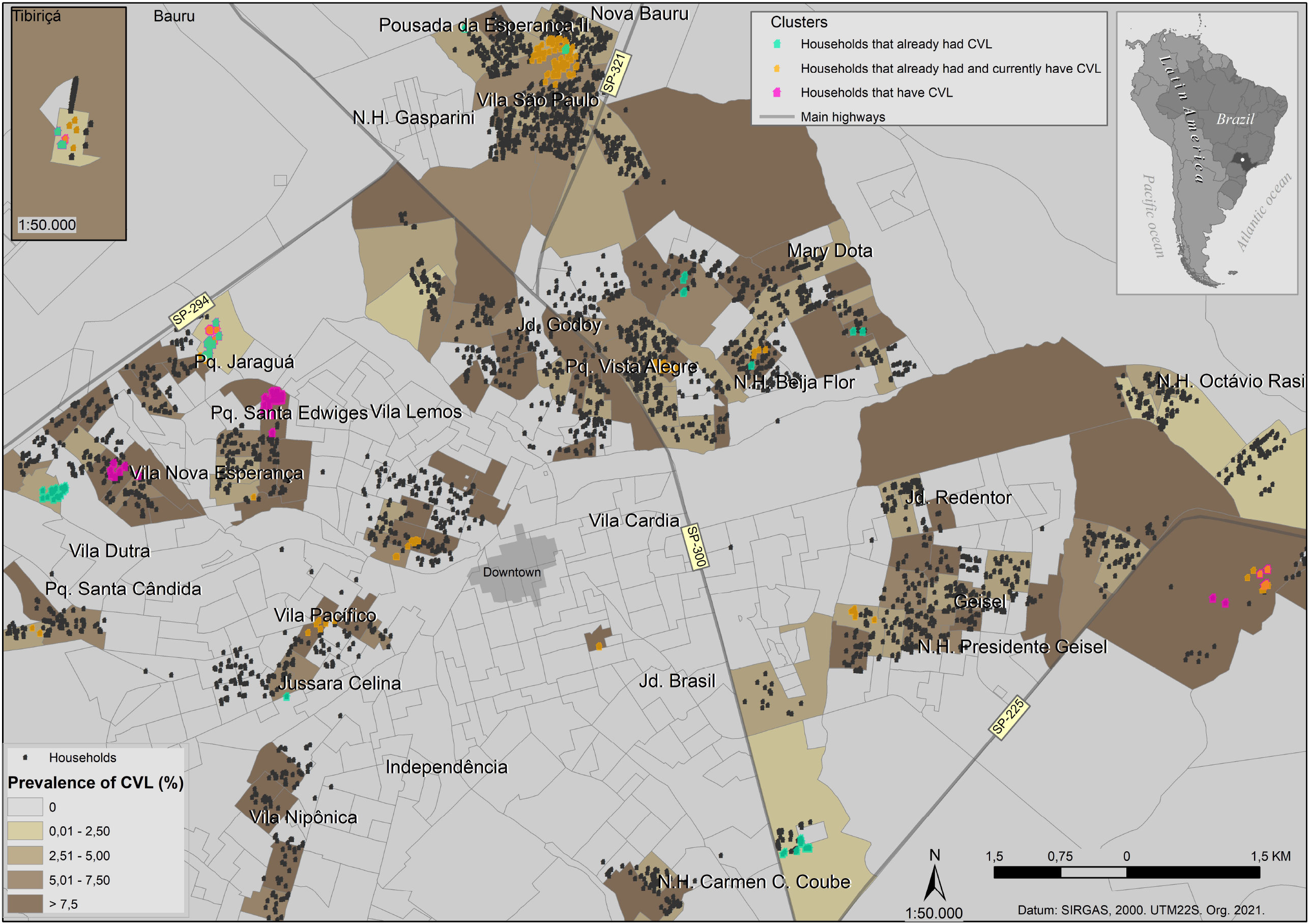
Geocoded households of Bauru, stratified by prevalence and cluster analysis. Each black dot is a household with no identified cluster. Magenta dots are the clusters of domiciles with infected dogs; green dots are the clusters of households that already had infected dogs; and orange dots represent the clusters of households that had and currently have infected dogs. Different size symbols and opacity households were set to ensure the spatial visualization of overlapped households.

The ratio of cases per sample concentration represented in the Kernel map shows high-risk areas in the Pq. Jaraguá, Pq. Santa Edwiges, and Vila Nipônica neighborhoods (Figure 4). Other high-risk concentrations in Kernel’s map represent the border effect.

**Figure 4:**
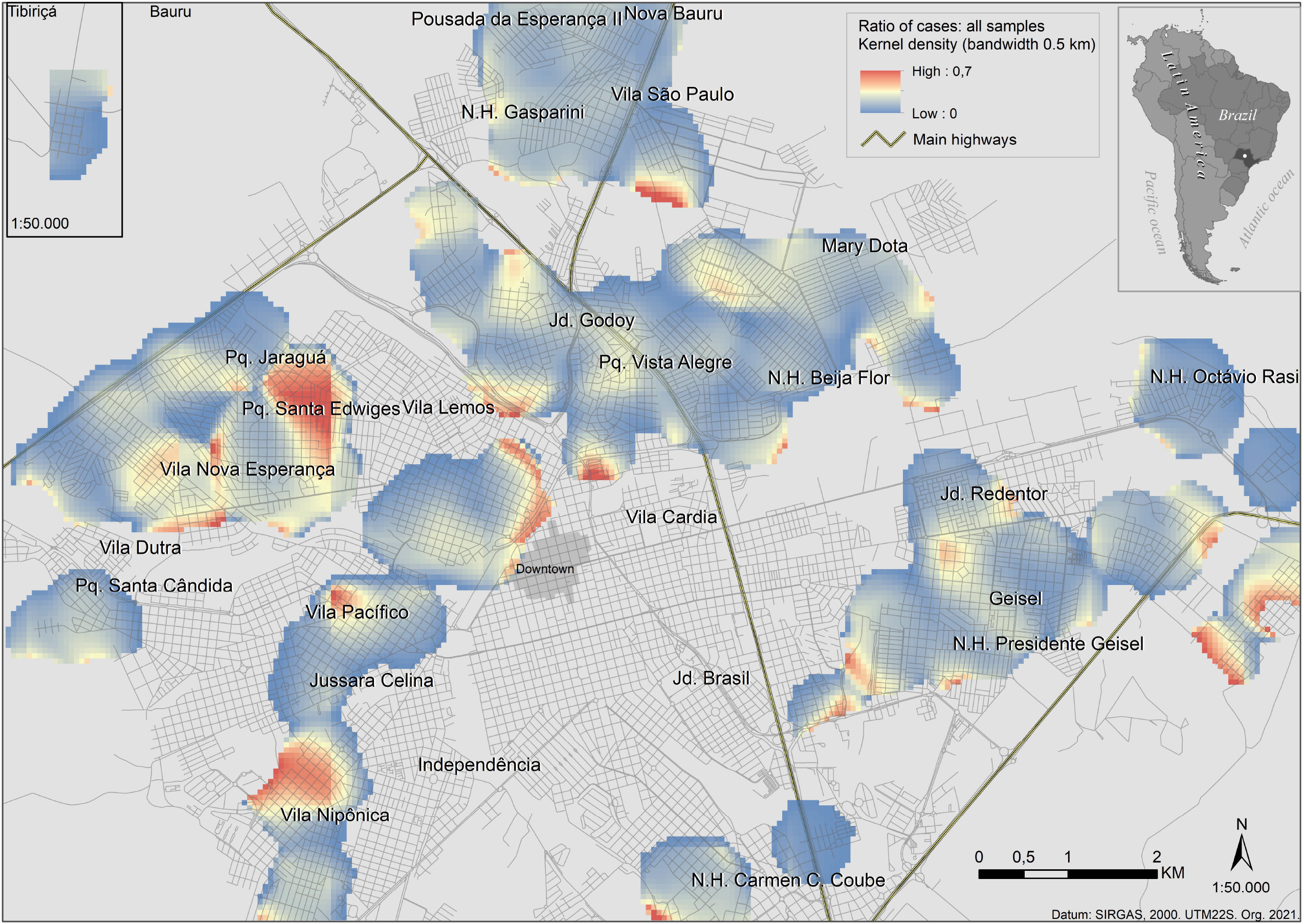
Kernel density for canine visceral leishmaniasis. Kernel density ratio map ranging from 0 (blue) to 0.7 (red), which gives a visualization of the risk diving the concentration of cases of CVL (Figure 4S) by the concentration of dog samples (Figure 5S). The areas of higher risk are in the west and southwest.

### 3.2. Pearson correlation

Pearson’s correlation was positive and moderate considering the number of infected dogs and the households investigated (0.508, p-value=0.000); positive low for infected dogs and the households that already had an infected dog/dogs (0.240, p-value=0.000); and positive and low for infected dogs and the households that already had and currently have a dog/dogs with VL (0.129, p-value=0.000). All conditions were statistically significant.

### 3.3. Binary logistic regression for visceral leishmaniasis

According to Table 1, 3,916 households were investigated, in which 16.7% (656/3,916) already had a positive dog - independently when it was. Nowadays, 8,7% (341/3,916) of the households have at least one positive dog. From the households with a positive dog/dogs in the past, 17.0% (112/656) still have a positive dog/dogs currently.

The maximum number of dogs per household was 17, the mean was (*m* = 1.67), with the standard deviation of SD = 1.08. In an area of influence of a household (*a* = 31,374m2), the maximum number of dogs was 58, the mean was *m* = 16, and the standard deviation *SD* = 11.37. Households with only one dog represent almost 60% of the domiciles, households with two dogs 27.6%, and households with more than three dogs 14%.

The odds ratio (OR) for CVL increased proportionally to the number of dogs (Table 2). The OR for number of dogs examinaded was 1.37. For households with only one dog was 0.40 and increased 242% for those with two dogs (OR=1.39); and 97% when more than two dogs (OR=2.70). For households that already had a positive dog, the OR was 2.73. OR for the area of influence of household (buffer of 100m) also increased according to the number of dogs. From 10 to 20 dogs, OR was 1.25 and increased 120% for 21 to 30 dogs (OR=2.76). The influence area of household with more than 30 dogs increased more than 150% (OR>7). In an area of influence of household, households that already had a positive dog/dogs with VL increase the chances 299% (OR=2.99), similarly to the analysis of the households that already had dog/dogs with VL (OR=2.73).

**Table 2:** Binary logistic regression of Canine Visceral Leishmaniasis diagnostic, dog count in the buffer zone extraction versus the number of positive dogs and human cases in Bauru

|  | Description | Odds Ratio | (95% CI) | P-value |
| --- | --- | --- | --- | --- |
| *Household and CVL* | Already had a dog/dogs with VL | 2.73 | 2.14 - 3.48 | 0.000* |
|  | 1 dog/household | 0.40 | 0.32 - 0.50 | 0.000* |
|  | 2 dogs/household | 1.39 | 1.10 - 1.76 | 0.006* |
|  | >2 dogs/household | 2.70 | 2.09 - 3.48 | 0.000* |
|  | Number of examined dogs | 1.37 | 1.27 - 1.48 | 0.000* |
| *Area of influence of household (buffer of 100m) and CVL* | Already had a dog/dogs with VL | 2.99 | 2.60 - 3.44 | 0.000* |
|  | Number of examined dogs | 1.10 | 1.09 - 1.11 | 0.000* |
|  | ≤10 dogs | 0.23 | 0.20 - 0.26 | 0.000* |
|  | > 10 ≤ 20 dogs | 1.25 | 1.10 - 1.42 | 0.001* |
|  | >20 ≤ 30 dogs | 2.76 | 2,28 - 3,35 | 0.000* |
|  | >30 ≤ 40 dogs | 7.73 | 5,25 - 11,39 | 0.000* |
|  | >40 ≤ 58 dogs | 7.29 | 4,69 - 11,34 | 0.000* |
| *Area of influence of household (buffer of 100m) and human cases* | Number of positive dogs | 1.16 | 1.09 - 1.24 | 0.000* |
|  | Number of samples | 1.02 | 1.01 - 1.02 | 0.000* |
|  | ≤10 dogs | 0.75 | 0.64 - 0.88 | 0.000* |
|  | > 10 ≤ 20 dogs | 1.16 | 1.00 - 1.35 | 0.053 |
|  | >20 ≤ 30 dogs | 0.77 | 0.62 - 0.96 | 0.022* |
|  | >30 ≤ 40 dogs | 1.27 | 0.95 - 1.70 | 0.104 |
|  | >40 ≤ 58 dogs | 2.61 | 1,93 - 3,53 | 0.000* |

Dependent variables are CVL and HVL cases. Explanatory variables are the number of dogs, infected dogs, and the condition of the households that already had an infected dog/dogs. \*statistical significance.

Considering the human cases in an area of influence of household, the number of dogs increased the chances for HVL 102%, and the number of positive dogs 116%, demonstrating the association between canine and human VL. The number of dogs increased the chances 261% for more than 40 dogs.

### 3.4. Spatial risk

Considering high OR for CVL according to the number of dogs, we created the spatial models using the number of dogs as a predictor. Figure 5 shows that both models (geostatistical and GAM) were considerable commonality in the spatial pattern. Higher risk is in the borders of the city, especially in the northwest and in the southeast. The last one can be a border effect. Moreover, both models are consistent with the Kernel density ratio map (Figure 4).

**Figure 5:**
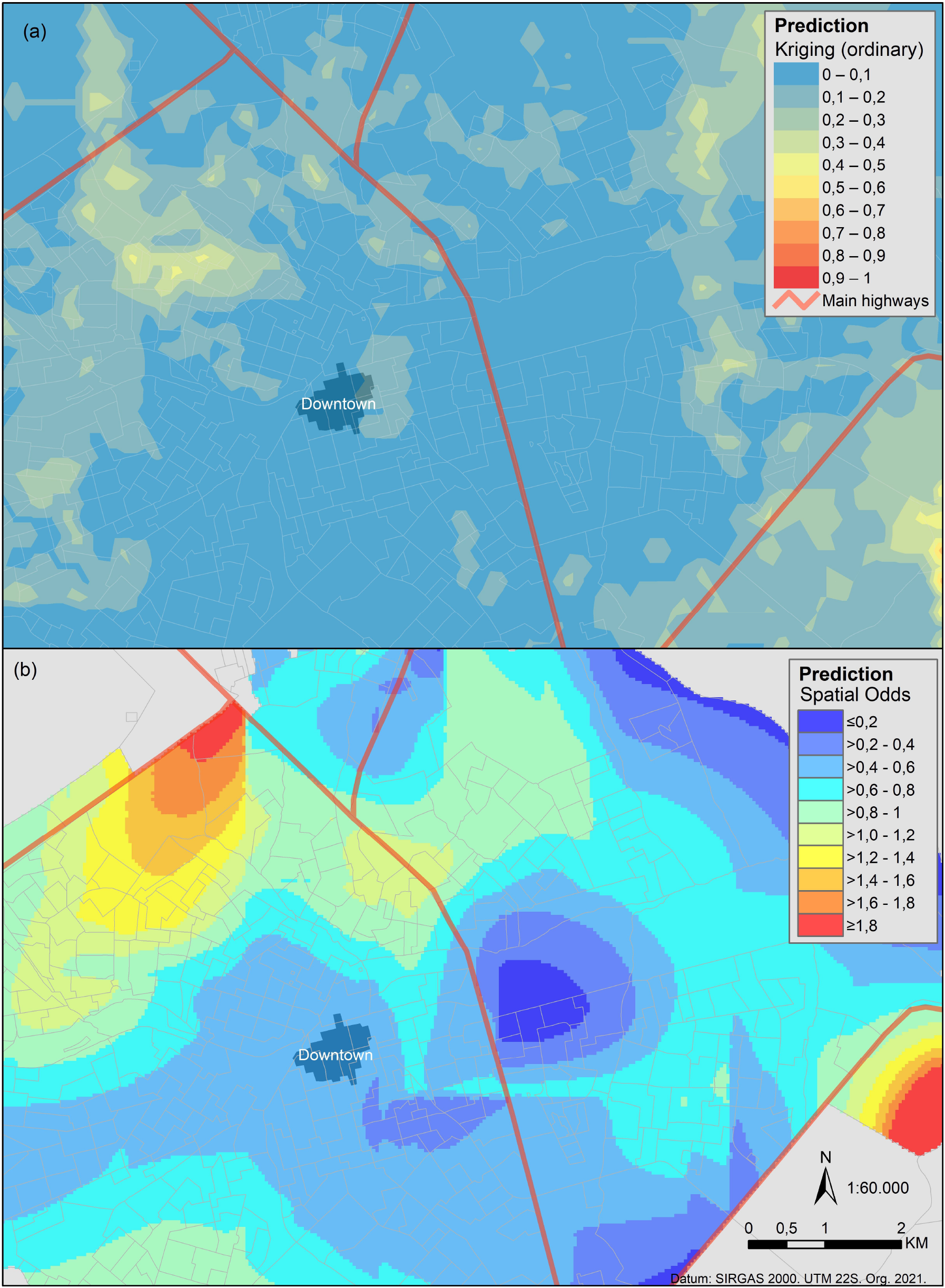
predicting the risk for canine visceral leishmaniasis using geospatial methods. Spatial prediction of CVL based on the number of dogs. Risk is scaled from low (blue) to high (red), as shown by the legends. (a) Geostatistical approach using the ordinary Kriging method. (b) Generalized additive model.

### 3.5. Cross-validation

Kernel, Geostatistical and GAM models were plotted in the ROC graph (Figure 6). The Kernel ratio presented the best threshold of 0.059, a sensitivity of 88%, a specificity of 62% and an accuracy of 64%. Geostatistical model presented the best threshold of 0.075, a sensitivity of 65%, a specificity of 52%, and an accuracy of 53%. GAM model presented the best threshold of 0.076, a sensitivity of 88%, a specificity of 19%, and an accuracy of 25%. The first had an AUC of 0.81 (CI 0.76 - 0.85), the second of 0.59 (CI 0.53 - 0.66) and the third of 0.54 (CI 0.47 - 0.60). The Kernel density ratio map presented the best performance in the ROC graph.

**Graph 1:**
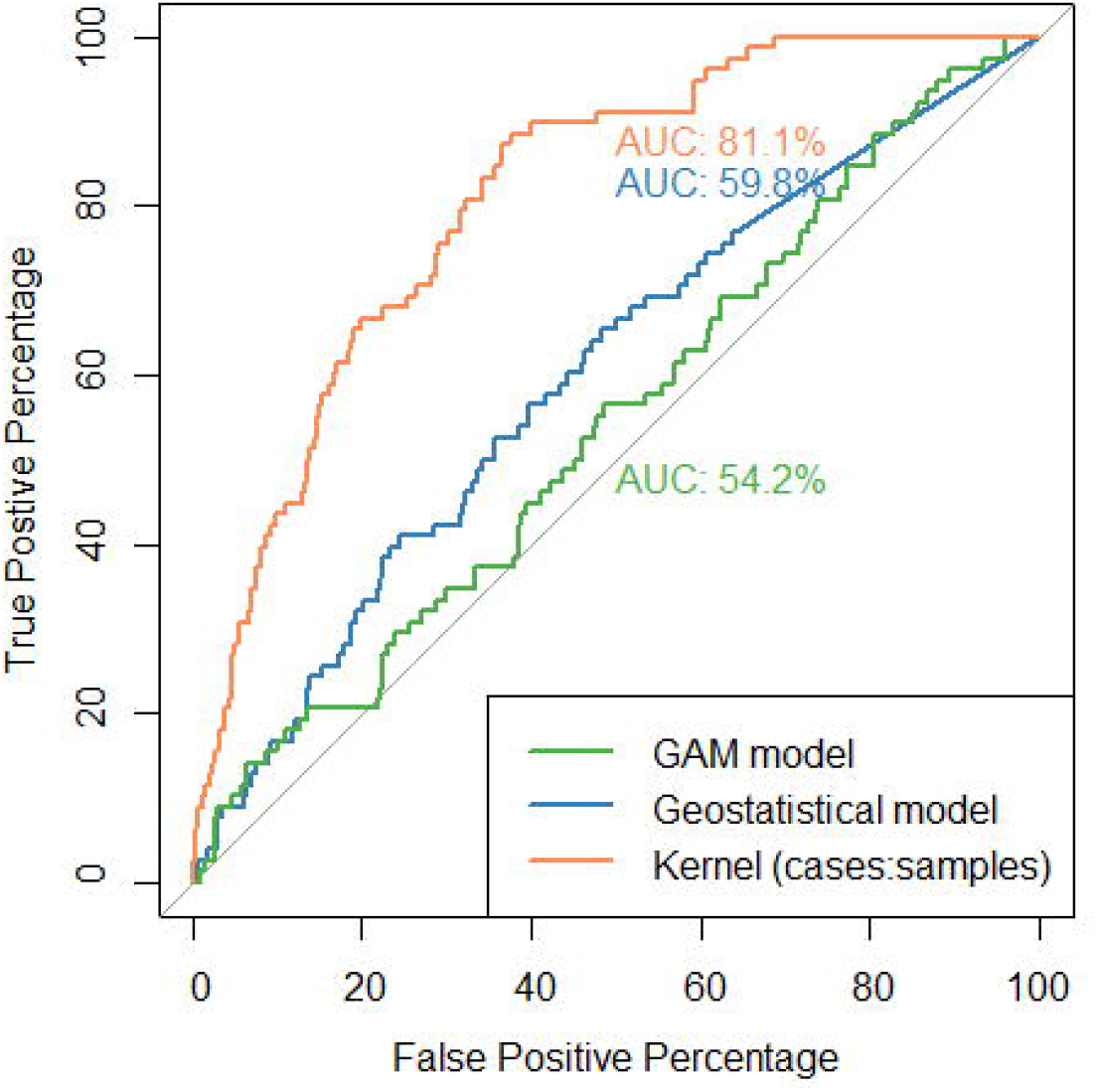
Area under the receiver operating characteristic (ROC) for canine visceral leishmaniasis. For each model, the AUC was calculated with 95% confidence intervals. The best model in predicting canine risk disease was the Kernel density ratio map.

## 4. Discussion

In the current study, we found a CVL TR DPP® sero-reaction rate of 8.1% (535/6578) and 5.6% in both TR DPP® and ELISA diagnoses (369/6578), likely consistent with an endemic area of Sao Paulo State, Araçatuba, where the average prevalence between 2010 and 2015 was 6.8% (26). Studies in other states of Brazil found a variable range of values of prevalence, for instance, 4.38% in Londrina, Paraná (27); 8.1% in the communities of Cuiabá river(28), and 19.2% in Rondonópolis(29), both in the State of Mato Grosso; 4.16% in Belo Horizonte, State of Minas Gerais (30); and a high prevalence of 50.3% in Buerarema, State of Bahia (31). In Bauru, we found different spatial prevalences, ranging from 0 to 50%. Lamattina et al. (2019) found prevalences per site varying from 0 to 80% (32) and Carvalho et al. (2018) from 0 to 35%(29).

Particularly, prevalence can reveal bias once it may not represent the real number of canines. Overall, the serosurveys are directed to human case areas and/or areas of a suspect or identified canine VL case (19)(8). Historically, in Bauru, some neighborhoods have never performed a serosurvey before this study. On the other hand, some neighborhoods were investigated more than once since the first human case appearance, recognized by its recurrence of CVL. Our study planned the serosurvey to collect a large number of dog samples in different neighborhoods, giving a panorama of VL’s endemicity and spatial epidemiological profile in a short time. Nevertheless, examined canines comprised less than 7% of the estimated dog population (6,578/99,815 dogs).

In the present study, our scale is the household instead of only the dogs, identifying spatial characteristics regarding the domiciles and canine population. We highlight that on the household scale, the positivity index of domiciles with infected dogs (8.7%) is superior that the global prevalence of CVL (5.6%), which emphasizes the importance of the household in the disease context. Clusters of households that already had CVL can point out the areas that remain a source of infection and are unnoticed. Almost all investigated areas had these clusters. Additionally, asymptomatic dogs can be highly competent (33) and remain a source of infection without being identified. They contribute to the silent endemic areas. It can turn out into highly endemic areas or possibly a human case site. Cluster gave us previous information of critical areas regards CVL.

The recent expansion of VL to new endemic areas has been attributed to the adaptation of *L. longipalpis* (sandflies) to naïve ecological niches. The risk of expansion of VL increases in areas identified as migratory poles of attraction. Moreover, CVL has been highlighted as the primary cause of outbreaks (34). In these areas, canine enzootic disease precedes the appearance of human cases. In our study, CVL increased the chances 102% for human cases and 116% for dogs, demonstrating an association between canine and human VL. Other studies found that the risk increased substantially for individuals when the presence of seropositive dogs (35) or previous cases of CVL in the household (36)(37). Furthermore, we identified a clustered pattern for both human and canine cases from approximately 500m.

As we identified, households with CVL and the dog population can increase the chances for VL and maintenance of natural foci of *Leishmania infantum* transmission to human and animal hosts, which urges specific public policies focused on animal health, especially in areas target as critical. We found the same mean number of dogs per household (1.6), as reported in previous research (18). Less than 15% of the investigated households have more than 3 dogs, which is the minority, easily to monitor and investigate as a possible infection site. In the neighborhoods where the canine population is large, animal health assistance is required. Therefore, canine population dynamics must be considered in public policies.

Our results revealed that the risk of increasing CVL or human cases oscillated by areas. Of note, kernel maps studies have used the total number of cases or applied a constant(30). Our study used the number of cases and samples, which gives a visualization of the risk. In accordance, (29) used the same methodology and found a similar pattern of critical areas in the city’s borders, a pattern that seems to be expected in small and medium-sized cities of similar urbanization process in low and middle-income countries where VL is endemic. The Kernel density ratio map was the best in the ROC graph, showing spatial analysis tools potential.

Spatial models predicting disease risk are promising for decision-making regarding the control of VL. Such studies use machine learning for cutaneous leishmaniasis vectors prediction (38) or human cases prediction (39). Bi et al. stress that future research about VL should focus on spatial simulation and agent-based simulation(40). Machine learning is a novel approach that allows the forecast of disease risk. It can anticipate disease transmission dynamics and identify disease control strategies to fight endemic and emerging diseases (40). Our models bring new insights for thinking VL through canines from a social perspective, which has been one of the most debatable points of control programs and tends to be of low priority in the context of general public health.

It is a time of changing public policies in relation to VL. The general principles that guided the past control programs are now questionable. Brazilian VLCP, performed by municipal levels, have presented operational difficulties in executing VL control strategies(41). Additionally, we highlight the unavailability of the proven effectiveness of technical alternatives for laboratory diagnosis, identification, and elimination or protection of reservoirs (42).

In Brazilian cities, culling dogs has been recommended as a control measuring to reduce VL (19)(43), which creates a dog stigma. Culling dogs is highly controversial, considering the time between diagnosis and action; issues from animal protection societies; rapid replacement of euthanized dogs as well the entrance of new animals into the households (44)(45)(46); and the persistent disease spread. Even in the academic environment, culling dogs have been critically discussed (42,47)(48)(49). By contrast, other strategies, such as the use of dog collar with insecticides for sand flies, or vaccination (50)(51)(52), have been highly encouraged due to their effectiveness in reducing the population of vector, parasitic load, and potentially the VL transmission (53)(54)(55)(56).

Vast territorial areas should be treated by priority order, emphasizing different profiles of VL. Furthermore, considering the genetic diversity of vectors (57)(58) and the protozoa (59)(60), even at the local levels, seem to be alternatives to rethink new VL approaches. The decision-making should be supported by an integrated approach, considering education, health, and environment, including vectors, causal agent, canines, households, population density, urbanization, presence of buildings, industries, and environmental factors, such as vegetation, water bodies, temperature, and precipitation. Animal health needs to be discussed in public policies without its stigma. Furthermore, VL should be addressed in the context of One Health (42).

To conclude, this paper had several limitations that should be recognized. Firstly, we had to use the census tract information based on the human population to calculate the canine population grid because of the lack of animal information. This could be solved with an updated canine census, hardly achieved in low and medium-income countries. Secondly, the performance of our spatial models had medium and low accuracy, although the critical areas being commonly similar to the Kernel ratio map of higher performance, which emphasizes a high chance that the classifier distinguishes the positive class values from the negative. The better performance of the models could be improved with an updated census and adding real-world covariates when data become available. There are still research gaps concerning VL, and many areas of study remain unexplored. It remains the question of balancing the effectiveness and costs involved in such a VL control plan (40). As future work, the next step of our research is to analyze the canines’ role with new insights of controlling VL, for instance, canine cohort studies of insecticide-impregnated collars, vaccination, and treatment in different areas of this endemic site, as an individual and collective measure in the environment.

## 5. Conclusions

As a rule of thumb, one can say that the number of canines and the households impact the risk for maintenance of natural foci of *Leishmania infantum* transmission to human and animal hosts in endemic areas for VL. Overall, this study serves as a case study for regional and global applications. It reveals the importance of canines on the household scale in low and middle-income countries. It is time for changing VL public policies using a targeted plan of priority through spatial analysis. This statement invites further investigations regarding VL characteristics involving socioeconomic and environmental variables and VL in one health context.

## Supporting information

https://www.dropbox.com/s/xvgf0zhfg5kehkg/Supplem_mat.pdf?dl=0

## Data Availability

Data cannot be shared publicly because of ethics to preserve the privacy of people s address (location of the household) of spatial data. Data can be requested in CVE and or Center for zoonoses control.

## Abbreviations

API: application programming interface
CVE: epidemiological surveillance center
CVL: canine visceral leishmanisis
ELISA: Enzyme-linked immunosorbent assay
GAM: generalized additive model
GIS: geographic information system
HVL: human visceral leishmanisis
IDW: inverse distance weighted
OR: odds ratio
TR-DPP: rapid test dual-path plataform VL= visceral leishmaniasis
VLCP: Brazilian visceral leishmaniasis surveillance and control program

## Authors’ contributions

Conceptualization, P.S.S.M.; J.E.T.; methodology, P.S.S.M; R.M.H; V.B.R.P.; H.H.T.; V.M.C.; software, P.S.S.M.; validation, P.S.S.M.; E.S.F.; R.B.G.; formal analysis, P.S.S.M.; investigation, P.S.S.M.; V.B.R.P.; J.E.T.; resources, P.S.S.M.; J.E.T; data curation, P.S.S.M.; writing—original draft preparation, P.S.S.M.; J.E.T; E.S.F.; writing—review and editing, P.S.S.M.; R.M.H.; V.B.R.P.; V.M.C.; H.H.T.; J.E.R.B.; L.R.P.B.C.; E.S.F.; R.B.G.; J.E.T.; visualization, P.S.S.M; supervision, J.E.T.; project administration, J.E.T.; funding acquisition, J.E.T. All authors have read and agreed to the published version of the manuscript.

## Acknowledgments

We thank Center for Zoonoses Control staff: Aline Fernanda Peral Cano, Cláudia Cilene Barbosa Gomieri, Josiane Silva Cano, Maria Emília Bodini Santiago, Roldão Antônio Puci Neto; Adolfo Lutz Institute students and staff: Aghata Regina de Oliveira Alves Palmeira; Amanda Gonçalves Martins da Costa; Alessandra Ventura Santos, Luana Ribeiro Manzi, Naíra Ruiz Lenharo, Maria Cristina de Carvalho; Graphical abstract was created with BioRender.com.

## Ethical statement

This study was approved under number 03/2000 in Ethics Committee on the Use of Animals in Research at the Adolfo Lutz Institute, Sao Paulo-SP, Brazil.

---

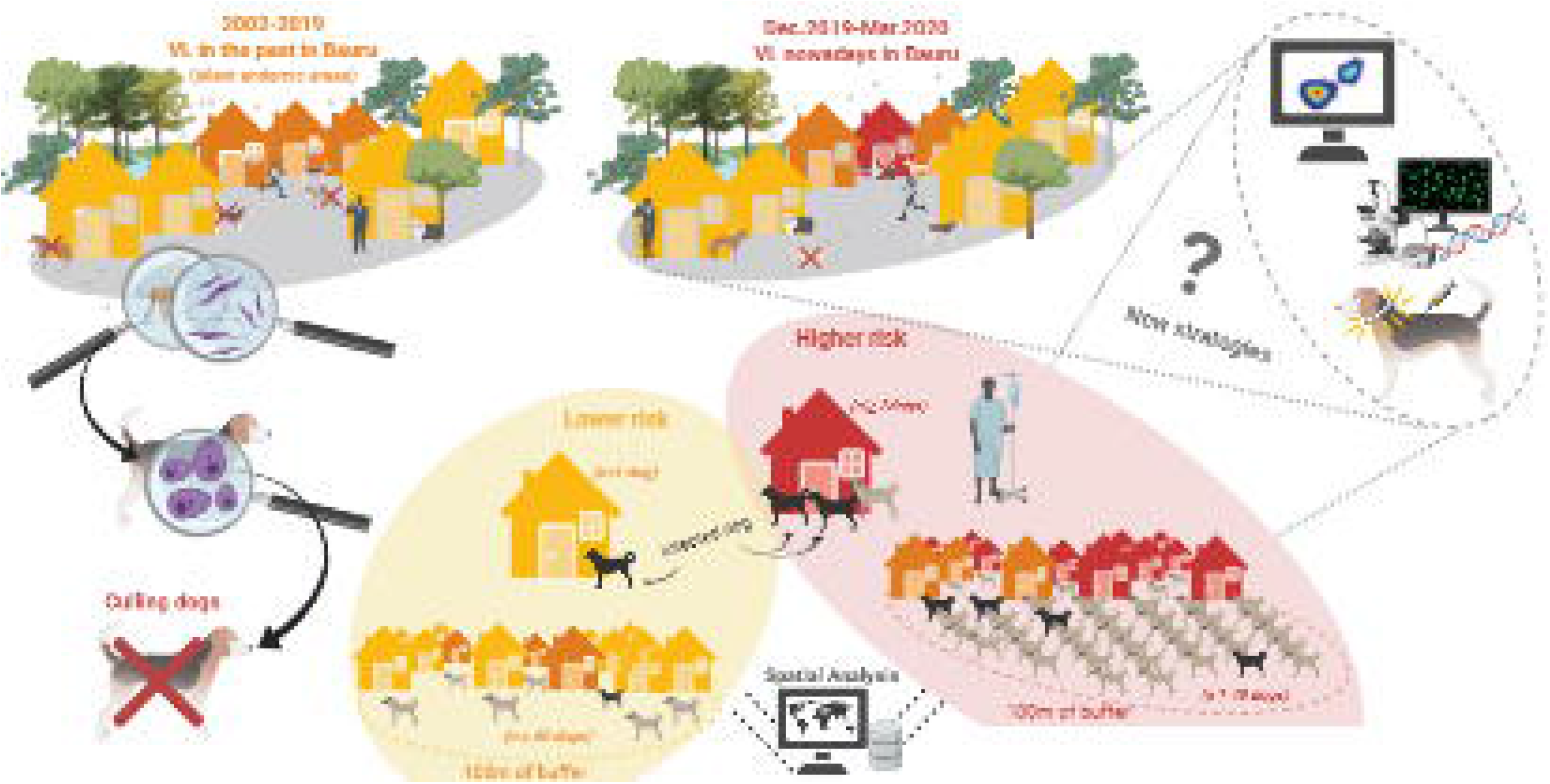

