## Supplementary material for "Impact of the household environment risk for maintenance of natural foci of *Leishmania infantum* transmission to human and animal hosts in endemic areas for visceral leishmaniasis in Sao Paulo State, Brazil": https://www.dropbox.com/s/xvgf0zhfg5kehkg/Supplem_mat.pdf?dl=0

*Figure 1S:* Synthesis of the Material and Methods.

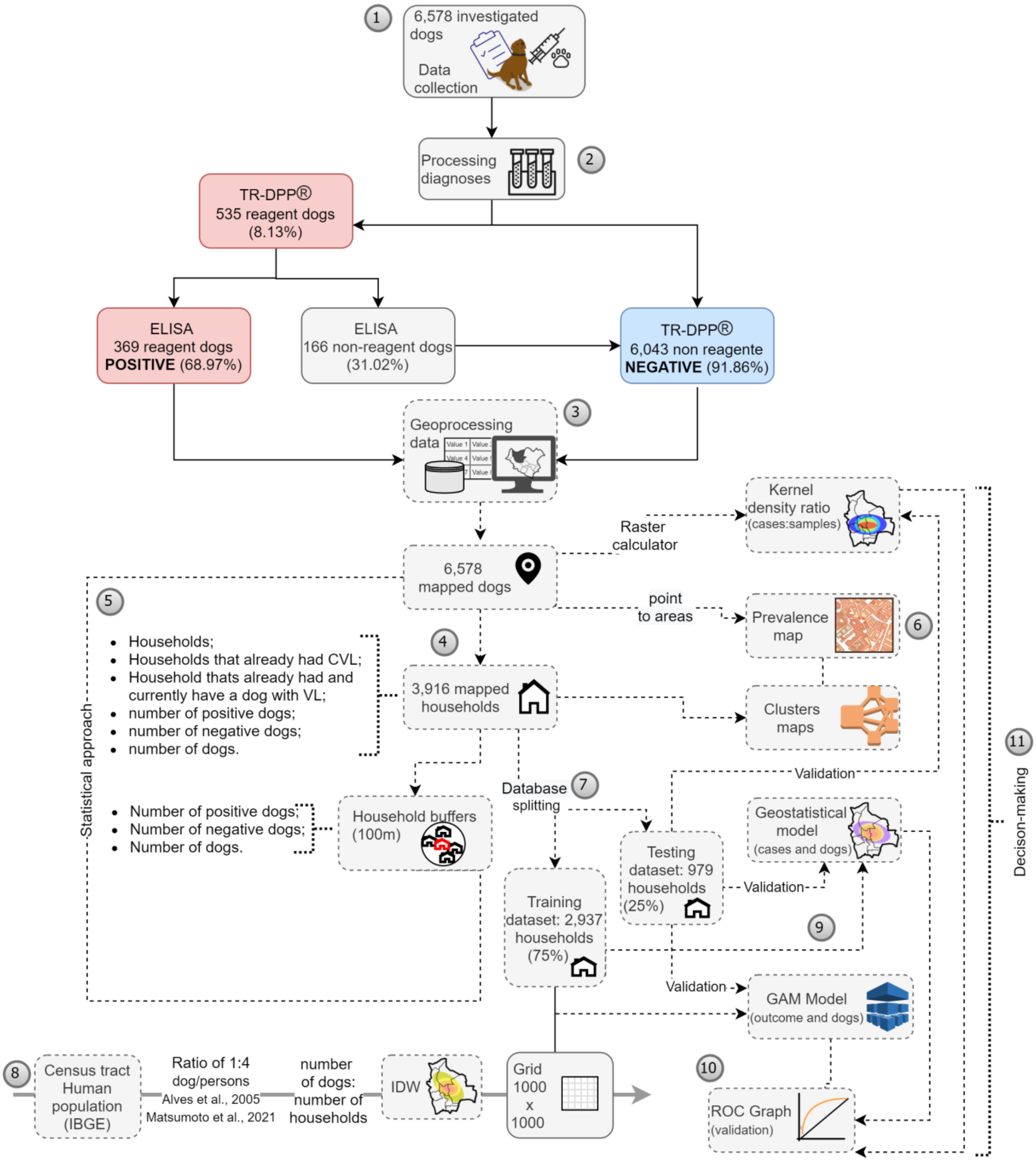


(1)The canine serosurvey was conducted from Dec.2019 to Mar.2021. Agents of the Center for Zoonoses Control visited 3,916 households and collected the blood samples of 6,578 canines. The applied survey is identified in *Figure 2S*. (2) In the laboratories, we tested the samples by TR DPP® and the reagents ones by ELISA. The non-reagent in RT-DPP® were considered negative according to the Brazilian Program of VL control. (3) All the dog samples were geocoded using an API of Google and manipulated in a GIS. (4) Spatial analysis tools were used to prepare data for thematic maps and statistical models. Households were mapped, and *buffers* (area of influence around the household) were created to calculate the number of dogs (positive or negative). Points (dogs) were transformed into areas (census tracts) to calculate the spatial prevalence. (5) Data were tabulated as binary, which allowed us to perform binary logistic regression models. (6) Thematic maps were created using spatial statistics. We calculated the dependence of cases at distances for households with CVL currently and for human cases of VL (Figure 3S), aiming to choose the bandwidth to calculate Kernel´s map. The map of the concentration of cases of CVL (*Figure 4S*) was divided by the map of concentration of samples (*Figure 5S*) to ensure the visualization of the risk (*Figure* 4). The prevalence map used the number of CVL cases divided by the number of collected samples in each census tract. Cluster maps were categorized according to having dogs with VL in the past and currently (*Figure 6S*). For each condition, it was exported the shapefile of only significant clusters. (7) For spatial models, the database was split into 75% (2,937 data) for training the models and 25% (979 data) for testing. A geostatistical model was performed considering CVL cases and the number of dogs in each household (*Figure 7S, Frame 1S*). We run a GAM model using a grid of 1000x1000 cells. (8) To estimate the number of canines in the grid, we used the relation of 1:4 (dog/persons) based on Alves et al. (2005) and Matsumoto et al 2021. The mean number of dogs was divided by the mean number of domiciles in each census tract to ensure the mean number of dogs in that point (centroid) (*Figure 8S*). We then create an interpolation (IDW) to calculate the mean number of dogs for all the grid (*Figure 9S*). The GAM model dataset presents the coordinates, the output (zero for non-cases and 1 for cases) and the number of dogs at each point. The best AIC was chosen to set the span function of the model (*Frame 2S*). (10) To conclude, data were validated using the testing data. We extracted the models' value of each testing dataset (at the same position of the coordinates). This allowed us to compare the observed versus expected values in each model, calculating sensitivity, specificity, and accuracy. A ROC graph was created to evaluate the performance of the models. (11) all the methodology and the results found here may be useful for decision-making regards public health.

*Figure 2S:* Survey applied in the collection of dog's blood.

| | | | | |
|---|---|---|---|---|
| Full name (guardian): | | | ID ____.____.____-___ | Cellphone: |
| Full address: | | | | |
| Dog's name: | ID (dog): | Breed: ☐ Poodle ☐ Yorkshir ☐ Boxer ☐ Dachshund<br>☐ Mixe ☐ Pinscher ☐ Shih Tzu ☐ Labrador ______ (other) | Symptoms ______ (other)<br>☐ sadness ☐ slimming | Obs.: |
| Sex:<br>☐ Female ☐ Male | Age:<br>☐ Puppy | Fur: ☐ Black-White ☐ White-Brown<br>☐ Black ☐ Light Brown ☐ Black-Brown | ☐ long nails ☐ wounds<br>☐ flaking ☐ conjunctivitis | |
| Wearing collar (insectiside)?<br>☐ No ☐ Yes | ☐ Young<br>☐ Adult | ☐ White ☐ Dark Brown ☐ White-Black-Brown<br>Result (DPP TR) ☐ Positive ☐ Negative ☐ Indeterminate | Had a dog with VL?<br>☐ Yes ☐ No | Agree to participate (cohorts)?<br>☐ Yes ☐ No |

The survey was applied at the moment of the collection of dog´s blood to detect anti-*leishmania* antibodies. It is a simple inquiry once the study covered a large number of dogs, and it was conducted by agents of the center for zoonoses control (not specialists). After the diagnose results, the guardians of the positive dogs were notified to schedule an appointment with a veterinarian.

*Figure 3S:* K-function for visceral leishmaniases at distances.

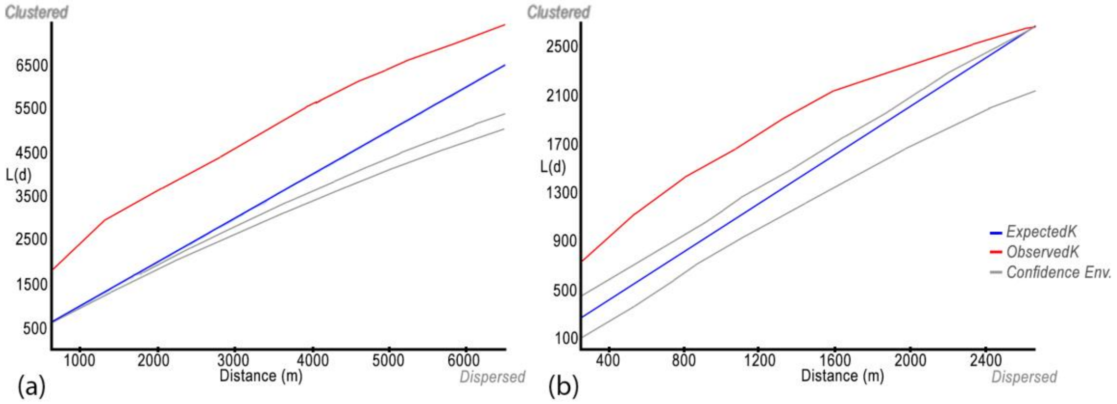


The red line is the observed values. The blue line is the expected for a random sample. Dashed lines represent the superior and inferior envelopes for statistical significance. (a) households with CVL currently; (b) human cases (2003-2019) of VL in Bauru, São Paulo, Brazil.

(a) (b)

*Figure 4S:* Kernel map for canine cases of visceral leishmaniasis.

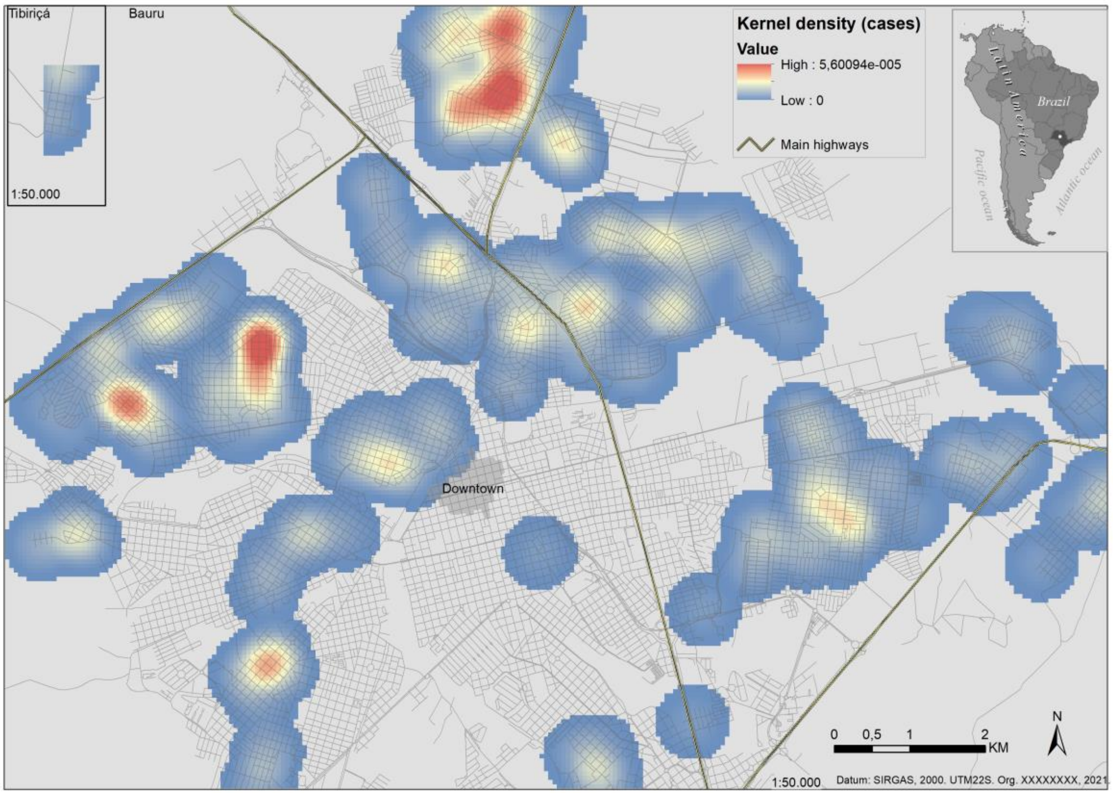


We performed a Kernel density map for the total number of cases using a bandwidth of 500m (approximately the minimal concentration of K-function). We select the default cells and the output in meters square.

*Figure 5S*: Kernel map for samples

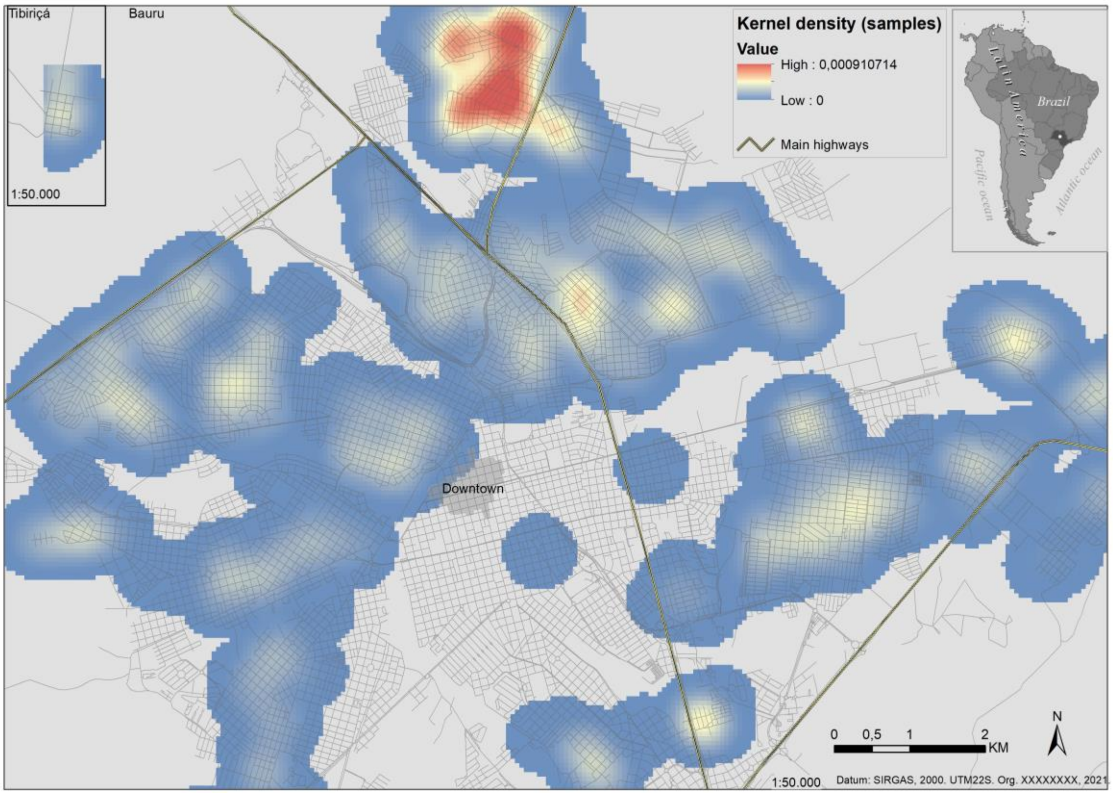


We performed a Kernel density map for the total number of samples using a bandwidth of 500m (approximately the minimal concentration of K-function). We select the default cells and the output in meters square.

*Figure 6S:* Cluster map for households that have CVL currently.

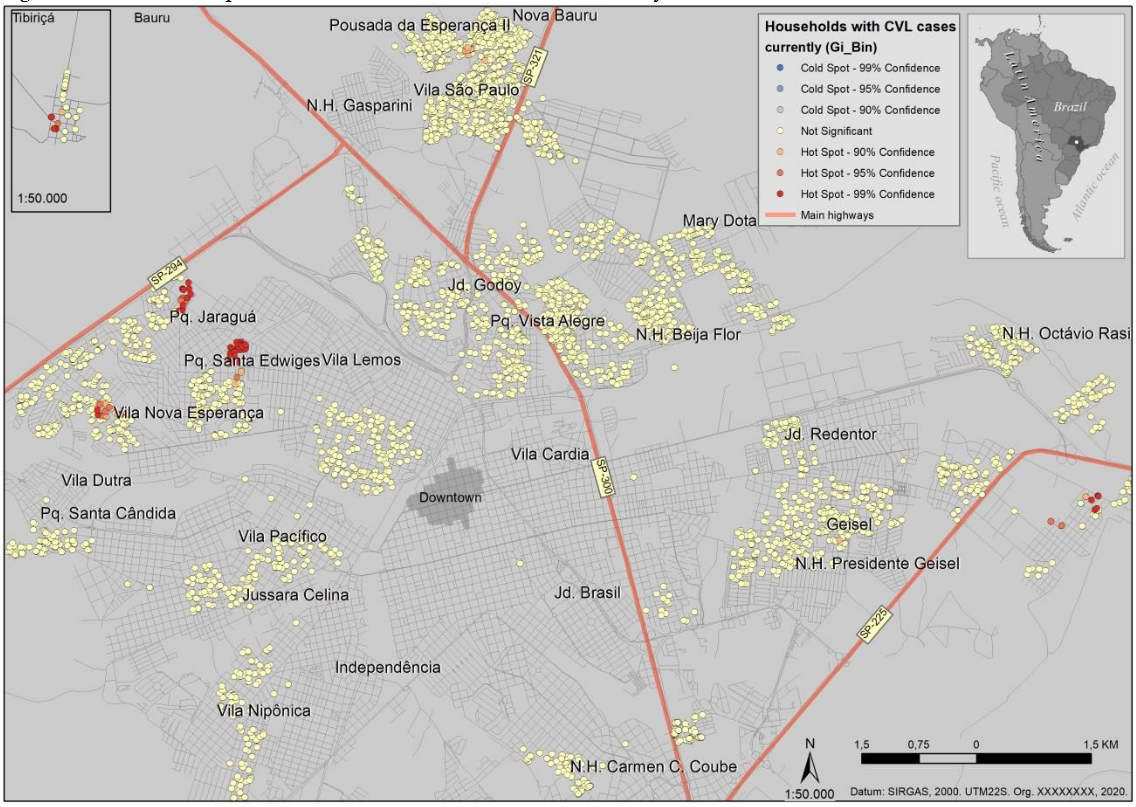


This is an example of a cluster map for the households that have CVL currently. For each category, cluster maps were created: i)households that have CVL; ii) households that already had CVL; iii) households that already had and currently have CVL. The coldspots and the non-significant data were excluded in the final cartographic representation (Figure 2).

*Figure 7S:* Semivariogram for canine visceral leishmaniasis and the number of canines.

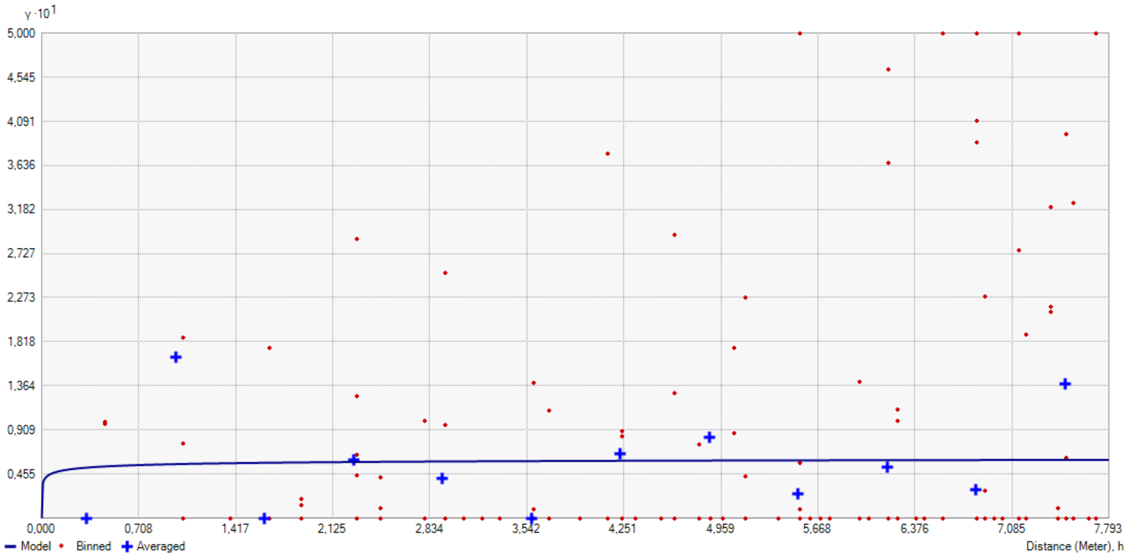


The stable theoretical model was adjusted to the points according to the parameters described below.

*Frame 1S:* Parameters of the semivariogram.

**Input datasets**
**-Dataset**
Type
Feature Class
Data field 1
LVC_PROJ
Records
2792

**-Dataset 2**
Type
Feature Class
Data field 1
Dogs
Records
2792

**-Method**
**CoKriging**
Type
Ordinary
Output type
Prediction
**-Dataset #**
1
Trend type
None
**-Searching neighborhood**
Standard
Neighbors to include
5
Include at least
2
Sector type
Four and 45 degree
Major semiaxis
182.670648783572
Minor semiaxis
182.670648783572
Angle
0
**-Dataset #**
2
Trend type
None
**-Searching neighborhood**
Standard
Neighbors to include
5
Include at least
2
Sector type
Four and 45 degree
Major semiaxis
182.670648783572
Minor semiaxis
182.670648783572
Angle
0
**-Variogram**
[ Semivariogram; Semivariogram ]
Number of lags
12
Lag size
0.649446959339
Nugget
[ 0; 0.465525660772 ]
Measurement error %
[ 100; 100 ]
Shift
[ 0; 0; 0; 0 ]
**-Model type**
Stable
Parameter
0.2
Range
3.8974342
Anisotropy
No
Partial sill
[ 0.062007611094; 0.045112598095; 0.045112598095; 0.660324964631 ]

We adjusted both datasets (CVL cases and the number of dogs) using a stable model.

*Figure 8S:* features of the grid of the GAM model

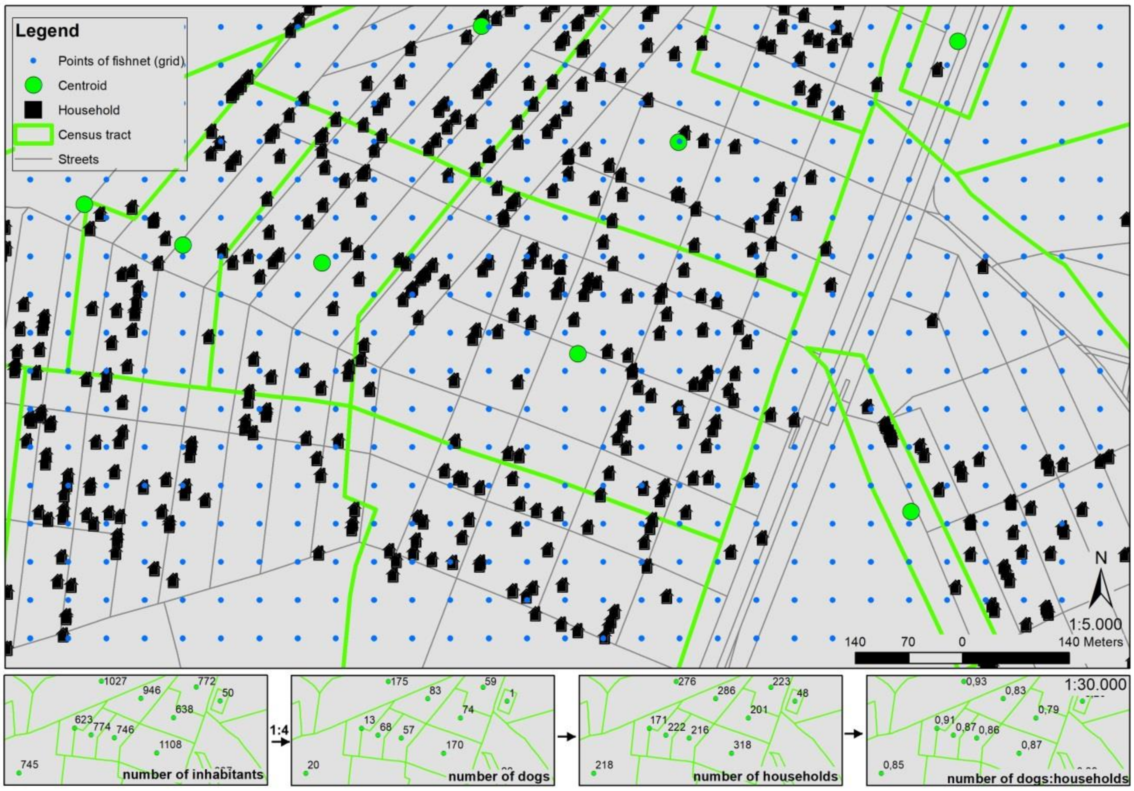


To calculate the number of dogs per domiciles, we used the study of Alves et al. 2005, an investigation conducted for São Paulo state cities, considering a ratio of 1:4 dogs/persons. We calculated the number of dogs based on the human population census tract (Matsumoto et al., 2021). We then used the number of households (IBGE,2010) to find the number of dogs at that point (centroid). Finally, a fishnet of 1000 cells versus 1000 cells was created to extract the point value of the number of dogs interpolation.

*Figure 9S:* Interpolation of the number of dogs using Iverse Distance Weighted (IDW).

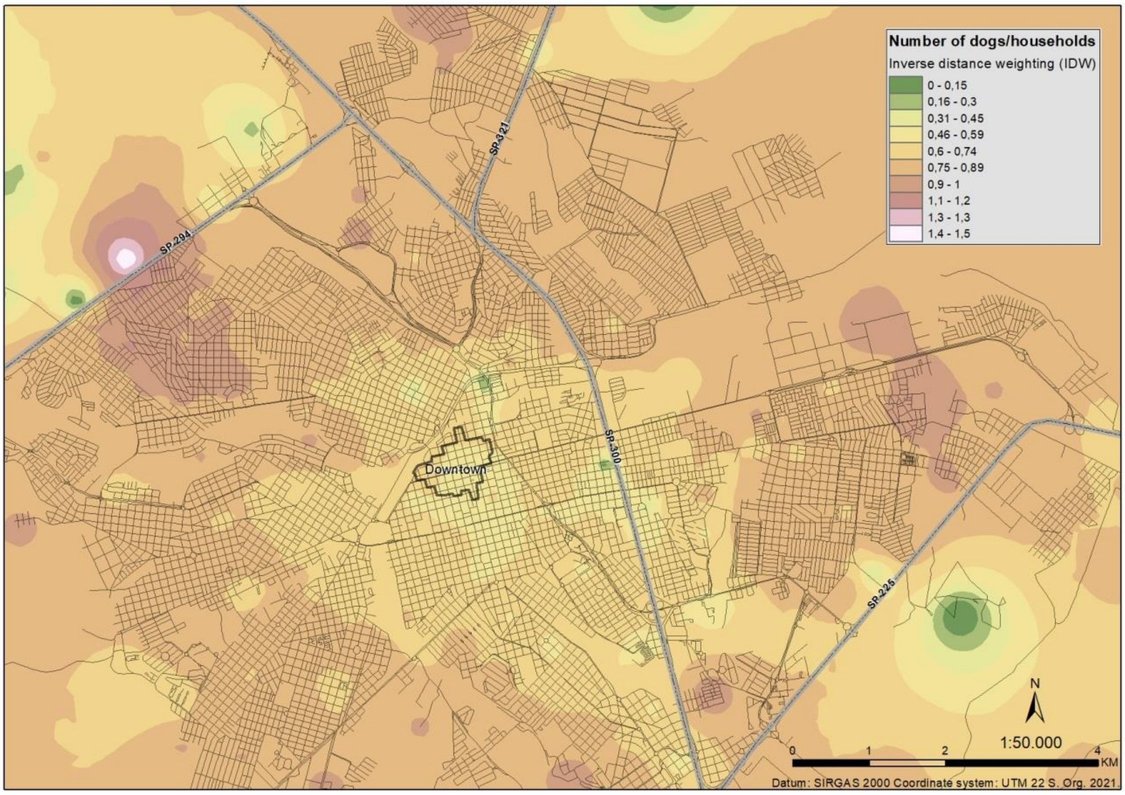


This method interpolates the estimative of the cell values using the average of the points in each region. We used the census tract data and the estimative of dogs according to Alves et al. 2005. The map shows a higher number of dogs per domicile (brown to white) in the city's outskirts and a lower number of dogs in the central areas (green to yellow). The grid (*Figure 7S*) extracted the IDW values of the correspondent location of each point. The grid can not see in the cartographic scale of 1:50000, but it is visible on the scale of 1:5000 (*Figure 7S*).

*Frame 2S:* AIC results for each span.

| Span | AIC training | Span | AIC training |
|---|---|---|---|
| 0.05 | 1749.292 | 0.55 | 1708.946 |
| 0.10 | 1709.256 | 0.60 | 1709.224 |
| **0.15** | **1704.05** | 0.65 | 1707.15 |
| 0.20 | 1705.583 | 0.70 | 1707.061 |
| 0.25 | 1709.209 | 0.75 | 1707.243 |
| 0.30 | 1709.771 | 0.80 | 1707.519 |
| 0.35 | 1710.728 | 0.85 | 1707.684 |
| 0.40 | 1710.429 | 0.90 | 1708.287 |
| 0.45 | 1710.122 | 0.95 | 1708.906 |
| 0.50 | 1709.284 | | |

We select the best (minimal) Akaike information criterion (AIC) for choosing the span function of our GAM model. The best AIC=1704, span=0.15.
